# *seqr* : a web-based analysis and collaboration tool for rare disease genomics

**DOI:** 10.1101/2021.10.27.21265326

**Authors:** Lynn S. Pais, Hana Snow, Ben Weisburd, Shifa Zhang, Samantha Baxter, Stephanie DiTroia, Emily O’Heir, Eleina England, Katherine Chao, Gabrielle Lemire, Ikeoluwa Osei-Owusu, Grace E. VanNoy, Michael Wilson, Kevin Nguyen, Harindra Arachchi, William Phu, Matthew Solomonson, Stacy Mano, Melanie O’Leary, Alysia Lovgren, Lawrence Babb, Christina Austin-Tse, Heidi L. Rehm, Daniel G. MacArthur, Anne O’Donnell-Luria

## Abstract

Exome and genome sequencing have become the tools of choice for rare disease diagnosis, leading to large amounts of data available for analyses. To identify causal variants in these datasets, powerful filtering and decision support tools that can be efficiently used by clinicians and researchers are required. To address this need, we developed *seqr* - an open source, web-based tool for family-based monogenic disease analysis that allows researchers to work collaboratively to search and annotate genomic callsets. To date, *seqr* is being used in several research pipelines and one clinical diagnostic lab. In our own experience through the Broad Institute Center for Mendelian Genomics, *seqr* has enabled analyses of over 10,000 families, supporting the diagnosis of more than 3,800 individuals with rare disease and discovery of over 300 novel disease genes. Here we describe a framework for genomic analysis in rare disease that leverages *seqr*’s capabilities for variant filtration, annotation, and causal variant identification, as well as support for research collaboration and data sharing. The *seqr* platform is available as open source software, allowing low-cost participation in rare disease research, and a community effort to support diagnosis and gene discovery in rare disease.

## 1. Introduction

Approximately 1 in 20 people worldwide are affected by a rare genetic condition, but approximately 65% of cases go undiagnosed due to limitations in diagnostic technology used, a lack of understanding of human genomic variation, and insufficient delineation of the mechanisms underlying disease (Chong et al., 2015; Boycott et al., 2017). Many of these unsolved cases move into the research realm, where exome and genome sequencing have been shown to increase the diagnostic yield by identifying complex variants and novel causes of disease (Clark et al., 2018; Palmer et al., 2021). With the large-scale uptake of exome and genome sequencing by research programs, including the USA’s Centers for Mendelian Genomics (CMG) (Chong et al., 2015) and Undiagnosed Disease Network (UDN) (Gahl et al., 2012), Canada’s Care4Rare program (care4rare.ca), and the UK’s Deciphering of Developmental Disorders (DDD) project (Wright et al., 2015), and Genomics England 100,000 Genomes study (Caulfield et al., 2017) among many others, vast and rapidly growing amounts of data are now available for analysis. To identify disease-causing variants in these large datasets, powerful filtering and decision support tools that can be easily accessed and used by researchers are needed.

Several existing tools have been developed in the academic sector for reviewing variants in defined gene lists (gene.iobio) (Di Sera et al., 2020) or for broader review and prioritization of variants in genomic analysis (Exomiser, GEMINI, PhenoDB, slivar) (Robinson et al., 2014; (Paila et al., 2013; Sobreira et al., 2015; Buske et al., 2013). This is also an area of development in the commercial arena, with many fee-based platforms available, such as those listed on ClinGen’s Genomic Analysis Software Platforms list

(https://clinicalgenome.org/tools/genomic-analysis-software-platform-list/). Each platform has its strengths, but none met all the needs of our team. Limitations included an inability to add specific features needed by our team for project management or analysis and inaccessibility to non-computationally experienced personnel. Additionally, proprietary software lacks features for broad data sharing and collaboration across the global rare disease research community, which is essential to uncover novel causes of rare genetic disease (Philippakis et al., 2015). Fee-based software can be cost-prohibitive for individual research groups, further limiting the utility of these platforms for rare disease research.

To meet the needs of the rare disease research community, our team developed *seqr* - an open-source, web-based platform for family-based analysis, data sharing, and project management. The National Institutes of Health (NIH) funded Broad Institute of MIT and Harvard Center for Mendelian Genomics (Broad CMG) has supported sequencing, analysis, and data sharing for an international collaborative network of rare disease researchers and clinicians since 2016 generating 23,166 exomes and 4,179 genomes from 16,803 families between 2016 and 2021. As the core analysis platform for the Broad CMG, *seqr* has enabled analyses of over 10,000 unsolved families, facilitated identifying a diagnosis for more than 3,800 of these families, and supported the discovery of over 300 novel disease genes. Current applications of *seqr* are primarily research focused, but *seqr* has also been incorporated in a clinical diagnostic pipeline at the Victorian Clinical Genetic Services laboratory in Melbourne, Australia. In this review, we describe *seqr’*s capabilities for rare disease genomics, and the technology stack underlying *seqr* that has facilitated scaling the system to analyze tens of thousands of exome and genome samples. A demonstration of the features described can be viewed on the Broad CMG website tutorials (https://cmg.broadinstitute.org/using-seqr).

## 2. Metadata storage, data upload, and project management functionality

An indispensable component of rare disease research programs is the need for affordable storage of large volumes of genomic datasets alongside easily accessible clinical information. A single platform with both functionalities streamlines the process of project management and analysis. The centralization of knowledge and data sharing across the teams that use the platform can also accelerate the pace of diagnosis and novel gene discovery.

The *seqr* platform possesses capabilities for data storage, case tracking, and data sharing. It allows for bulk upload of individual phenotype metadata in tsv, csv, or json formats for handling larger cohorts. It stores thousands of sample IDs with family pedigrees, phenotype information using the human phenotype ontology (HPO) format, additional sample metadata, and quality metrics. Data loading in *seqr* is optimized for a joint called variant call format (vcf) file from exome or genome data using the GATK pipeline, although joint-called vcfs generated with other callers have also been used successfully. Copy number variant (CNV) calls from exome or genome data generated by tools such as gCNV can also be loaded.

We are also currently working on integrating structural variant (SV) calls from genome datasets generated from GATK-SV (Collins et al., 2020). The user-friendly interface enables *seqr* to be used by a diverse team of variant analysts, researchers, clinicians, and project managers to manage sample data, review and prioritize variants, submit candidates to Matchmaker Exchange, and generate custom reports.

In *seqr*, an overview of a cohort can be accessed via the Project Page (Figure 2). Summary data about the project includes information about the families and data loaded in the project, stats on analysis status and MME submissions, along with gene lists used in the project and sub-cohort analysis of groups of similar cases. This is followed by a collapsed list of families in the project showing the analysis status, who has analyzed the case, if and when data was loaded, a high-level case description, and any saved variants. Additional detailed information about a case, including phenotype stored as HPO terms and sections for notes about analysis progress, can be entered on the Family Page (Supplementary Figure S1).

**Figure 1:**
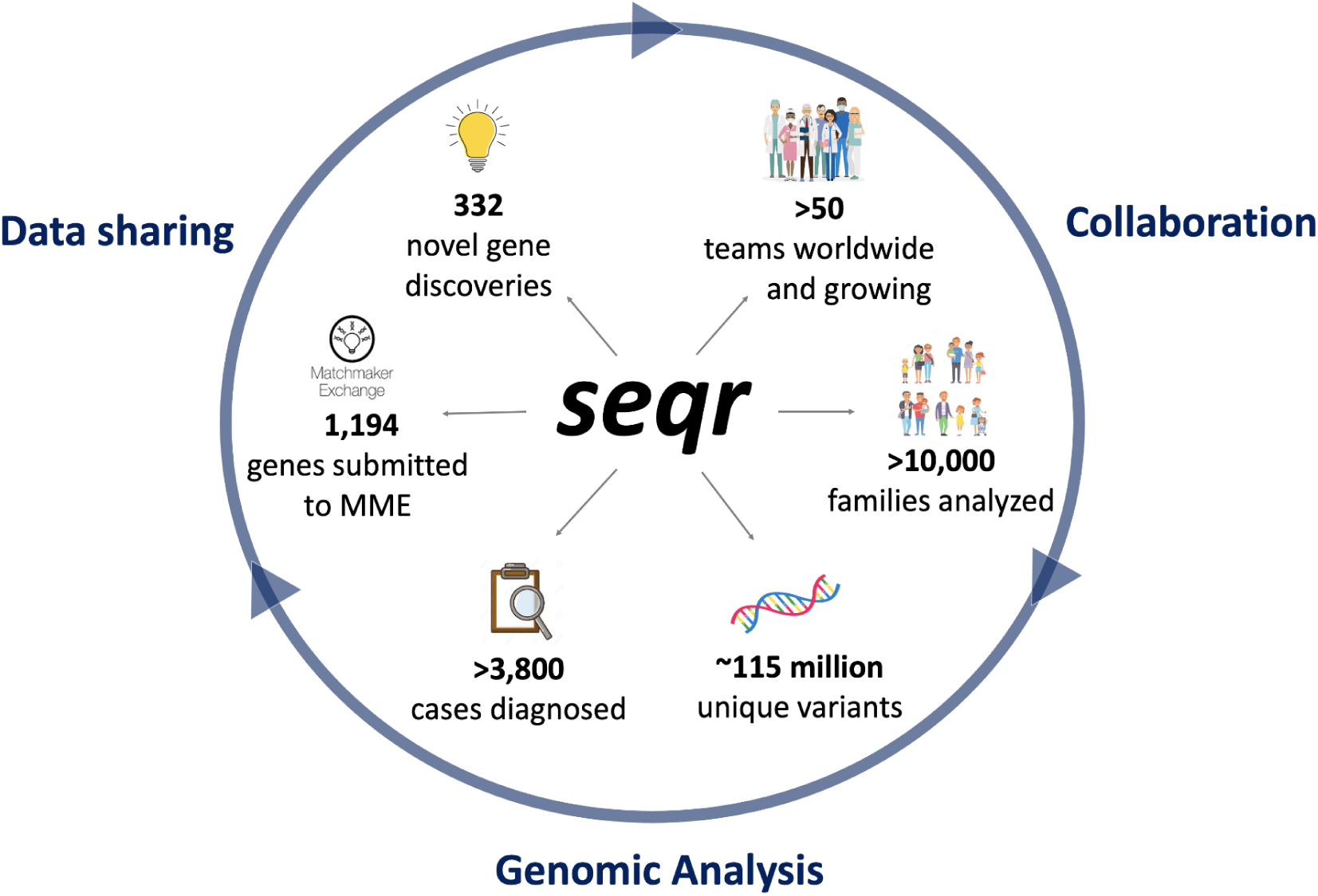
The *seqr* platform for family-based genomic analysis, research collaboration, and data sharing.

**Figure 2:**
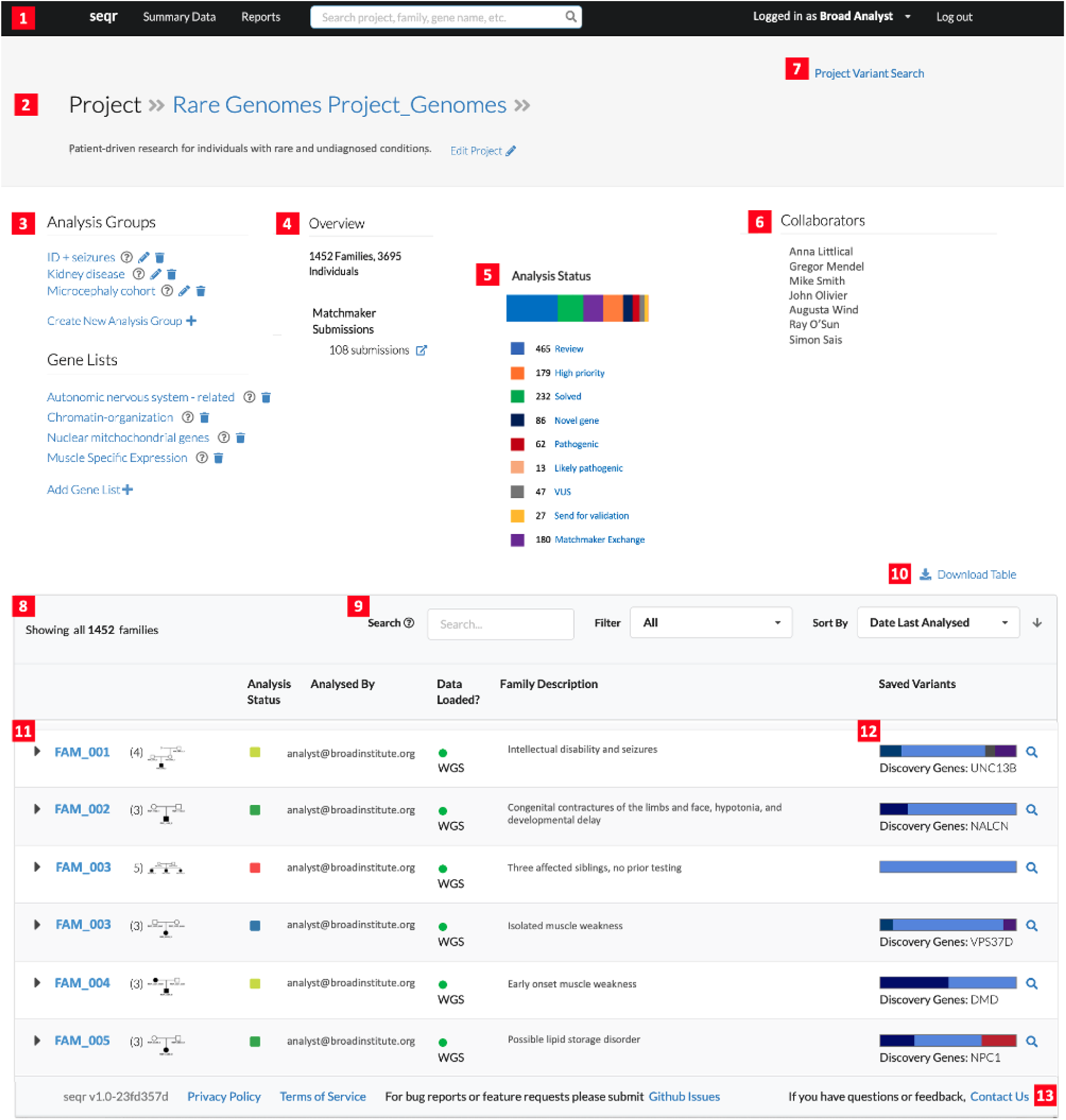
The Project Page in *seqr*, showing the Rare Genomes Project as an example. The top portion of the page contains (1) quick links to the main page and summary data; (2) project description details; (3) customizable analysis groups and gene lists; (4) project overview with high-level details of the number of families, type of data available, analysis status; (5) variants tagged in the project; (6) users with access to the project; (7) project-wide variant search function. The lower portion of the page lists the following functions or summary data (8) number of families in the project; (9) functions to search, filter, sort through families; (10) ability to download the list of families with analysis details; (11) overview of each family with the pedigree, details of the analysis status, the user who has analyzed the case, date the data was loaded, description of the family, and overview of the saved variants. Selecting the family ID opens the Family Page; (12) colored box highlighting tagged variants and quick link to the variant search page; (13) link to the GitHub repository for bug reports or feature requests, and the option to contact the *seqr* team.

## 3. Search/Filtration functionality

Variant searches can yield tens to hundreds of candidates for manual review, depending on the search parameters, type of sequence data, and the availability of familial data. Current clinical filtration workflows are largely based on published gene-disease associations, potentially excluding novel causes of genetic disease. Additionally, separate analysis of different data types (e.g. SNV/indel calls or SV calls) can limit the diagnostic rate due difficulties in recognizing compound heterozygous calls across variant classes.

The *seqr* variant search can efficiently filter SNVs/indels and SVs (from exome or genome data) in tandem. The search functionality is based on six core parameters including: suspected inheritance, reported pathogenicity in ClinVar and HGMD (optional feature depending on local licensing), type of variant, frequency in population databases, chromosomal region or gene list location (optional), and variant call quality. Search results can be further sorted by position, protein consequence, allele frequency, gene constraint, pathogenicity per ClinVar, gene-disease association per OMIM, and in silico scores. To enable researchers of various skill sets to participate in the filtration process, *seqr* has predefined de novo/dominant and recessive searches (the latter includes homozygous, compound heterozygous, and X-linked recessive in one search). If a suitable candidate has not been identified with these predefined searches, each search parameter can be customized, such as adjusting the inheritance filter to accommodate for incomplete penetrance in a family member, increasing the allele frequency to account for hypomorphic variants, or relaxing cutoffs for variant quality metrics to be more permissive. Customized searches can be saved for future use by a user.

The platform’s advanced search functionalities allow the creation of sub-cohort analysis groups such as for joint analysis of families with similar clinical presentations. Users can also focus variant searches based on user-created gene lists. Gene lists are encouraged to be entered as public lists to promote use by other teams using *seqr* for analysis. Sharing candidate disease genes can help other researchers to identify additional cases for inclusion in a case series publication. Publicly available gene lists maintained by the *seqr* team include curated assessments of the clinical validity of gene-disease relationships from PanelApp (Martin et al., 2019) and novel gene candidates from across the four Centers for Mendelian Genomics.

## 4. Analysis functionality

Assessing a subset of filtered variants for potential clinical significance requires comprehensive variant and gene level information. For each variant returned in a search, *seqr* displays in a single view the genotype, gene and Ensembl ID, chromosomal location, MANE transcript, disease association as per OMIM, missense and loss of function constraint metrics, variant quality scores, variant frequency in population databases and the callset, in silico scores, and ClinVar classification if available. While many systems include similar annotations, they are often displayed in a long series of columns that require more space and back-and-forth scrolling for review. By grouping and coloring data, using more horizontal space per variant, and providing additional information by hover over or with links, *seqr* shows a large amount of data for each variant that is quickly interpretable by an analyst. Variant quality assessment is made easy through the integrated genomics viewer (IGV) (Robinson et al., 2011) (Figure 4) for every variant returned in search, though use of this feature does require *seqr* to have the path and read access to the CRAM files. Filtration of SNVs/indels and SVs can be performed simultaneously, with results displayed together, to enable identification of compound heterozygous variation across CNVs/SVs and short variants (Supplementary Figure 2).

For genes with little or no evidence for disease, delineating the potential clinical significance of a variant is a laborious process of seeking gene-phenotype relationships by consulting numerous disassociated databases. To aggregate available data for analysis, *seqr* displays links to external resources, including gene-related information (PubMed, Monarch, DECIPHER) (McMurry et al., 2016; Shefchek et al., 2020; Firth et al., 2009), transcript information (gnomAD, GTEx) (Karczewski et al., 2020; GTEx Consortium, 2013), and functional animal model (MGI, IMPC) (Bult et al., 2019; Dickinson et al., 2016) databases (Supplementary Figure 3).

**Figure 3:**
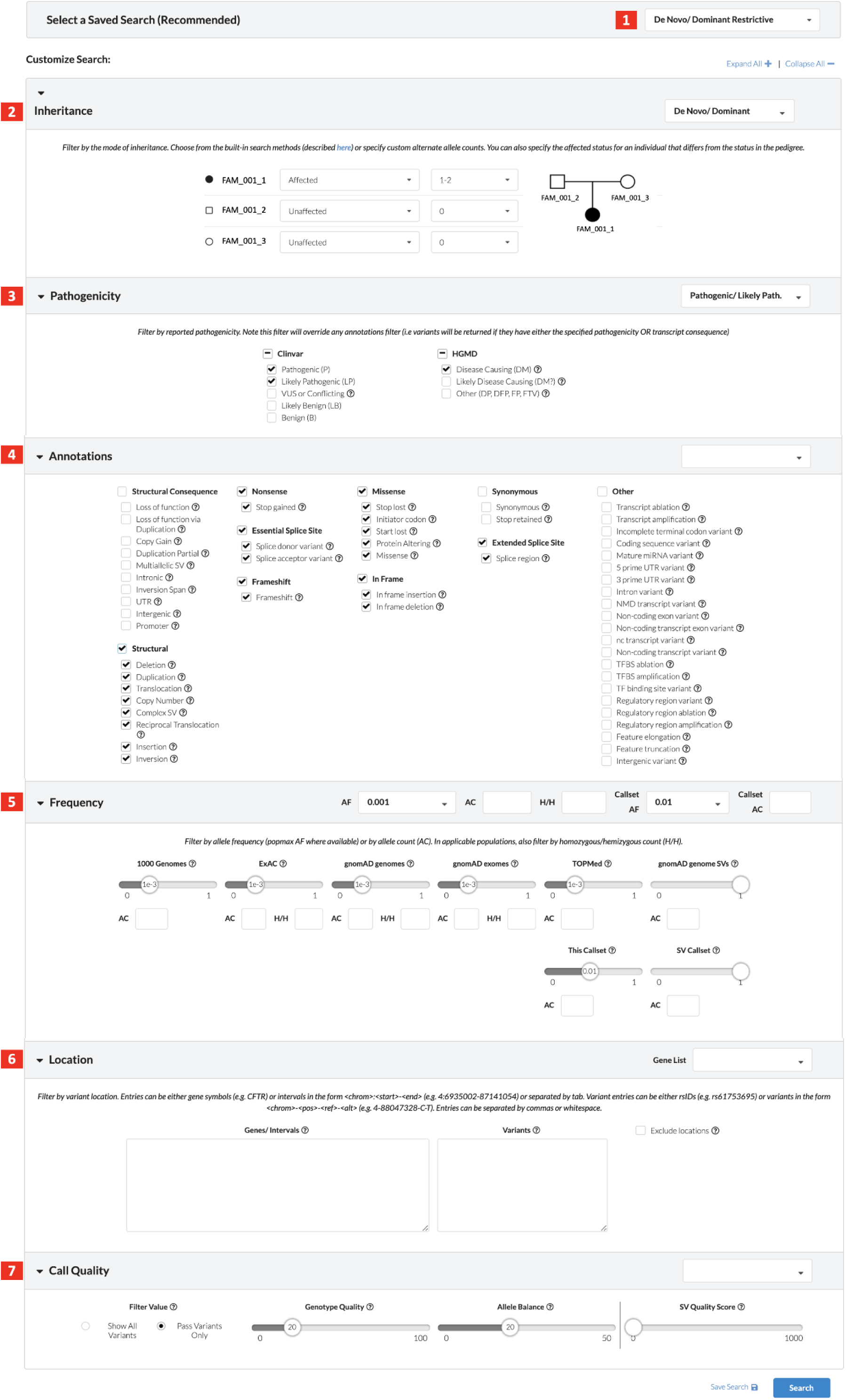
The Variant Search Page in *seqr* displaying settings for a de novo/dominant restrictive search from the (1) predefined searches with (2-7) options to customize each search parameter.

**Figure 4:**
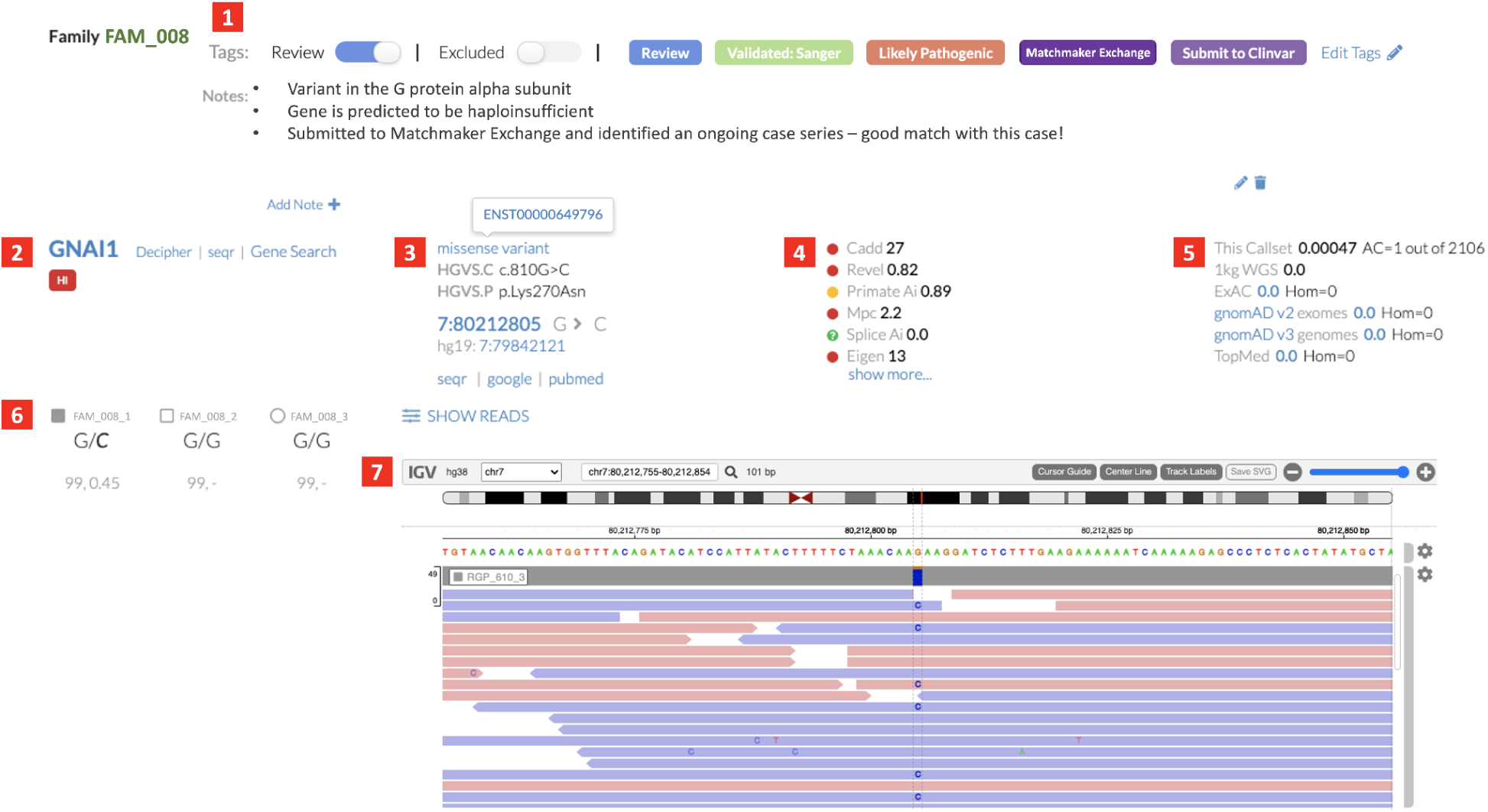
The variant view in *seqr* showing a *de novo* missense variant in the gene *GNAI1* as an example. (1) Tags have been used to mark the variant and additional notes have been added by an analyst; Variant information with (2) gene ID, link to the gene page in DECIPHER, *seqr* search function to display all previously saved variants in the gene, Gene Search for all variants in the gene present in any affected individual within this family; (3) variant HGVS nomenclature, with transcript ID, genomic location link to the UCSC browser, links to *seqr* search for this variant in the callset, Google and PubMed searches for the variant; (4) in silico scores; (5) variant allele frequency within the joint called vcf and reference population databases; (6) genotype for each individual with the genotype quality score and allele balance; (7) IGV read data in *seqr*.

Tracking variants as they proceed through the analysis pipeline can be challenging in large rare disease cohorts. In *seqr*, variant *tags* can be used to highlight variants of interest, marking it for further review, validation, or as the diagnostic variant. Users can filter for variants with a specific tag in a particular case or across a project such as those requiring orthogonal confirmation, classification of pathogenicity, or results ready for return to a participant. In addition to *tags*, the variant *notes* section can be used to record an analyst’s impressions and additional data gathered so that work is not duplicated across multiple searches on the same case. The public *notes* section is gene-specific and can be used to share information more broadly with all *seqr* users including relevant references, available functional studies, or case series in progress.

## 5. Data sharing

For many undiagnosed individuals with variants in genes with limited information, the presence of variants in the same gene in other cases can provide useful evidence for or against the role of the gene in disease. To enable data sharing among research labs and clinical centers across the globe, the Matchmaker Exchange (MME) (Philippakis et al., 2015) was developed to connect researchers and clinicians with cases that have variants in the same gene of interest. Through *seqr*, users can seamlessly submit a gene, variant, and HPO terms to MME. The *seqr* platform also makes contacting potential “matches” easy by auto-generating a customizable email (Supplementary Figure 4) with relevant variant and phenotype information listed in *seqr*. Gene submissions and contacts are recorded for easy tracking, and users can list additional notes regarding the gene submission such as matches or progress made. To date, seqr has aided in submitting and tracking >6,500 match communications.

## 6. Reporting

One of the biggest hurdles to rapid and comprehensive data sharing is the lack of structured data storage. Details about gene/variant evidence, data sharing activities, or patient phenotype are commonly stored in free text fields, which can be impossible to parse, and thus are difficult to include in data exports or other analysis reports without substantial manual effort. We address this issue in *seqr* by storing all accompanying metadata in discrete fields that can be used for creating data sharing files. By allowing our analysts and project managers to input this data into structured fields, rather than free text or attachments (e.g. PDFs), we can quickly meet the data model requirements for most genomic data sharing platforms.

In the case of the Broad CMG’s NIH progress reporting, we were able to substantially reduce the amount of time our team spent creating these semi-annual reports by transitioning from an entirely manual effort, which took weeks for our team to complete, to an automated report which takes minutes and can be generated at any time. Similarly, our CMG data is stored in NHGRI’s AnVIL (Analysis Visualization and Informatics Lab-space; https://anvilproject.org) and our AnVIL metadata reports, which include over 60 values varying from family and individual level data to sample and discovery details, can be generated by the project management staff on demand with minimal to no manual manipulation. The design of additional report formats takes development and software engineering time but allows for reproducible and expedient report generation. This infrastructure has allowed us to scale our data sharing activities without adding extra burden on our analysts and project managers.

## 7. Under the hood

The *seqr* platform is constructed as an open source project, designed to balance the needs of a small development team with those of a broad and ever growing user base. The underlying technologies were chosen for their maintainability and flexibility to allow their continued usage and evolution and also for their ease at being bundled and connected to streamline open source installations. Additionally, databases were selected and designed to both support *seqr’s* robust searching needs and ensure data integrity and stability.

The *seqr* web platform is built using a Python web framework to handle server-side functionality such as URL routing and database querying (see Supplementary Methods for details about the framework and databases). While *seqr* is designed to be platform-agnostic, it is currently hosted on Google Cloud Platform (GCP) and Kubernetes is used to manage deployment and resource management within GCP. Data storage in *seqr* is separated into two different databases: a read-only database which includes all annotated variant information, and a read/write database which includes all project and user metadata, including user-generated data like variant tags and notes. Additionally, *seqr* stores external reference data in a separate database. Such reference data includes gene level information from sources such as GENCODE (Frankish et al., 2019), OMIM (McKusick, 2007), and dbNSFP (Liu et al., 2020), as well as structured phenotype data from HPO (Köhler et al., 2021). This reference data is updated periodically to ensure accuracy but is not editable by *seqr* application users. Databases are backed up at regular intervals, conforming to best practices and FISMA requirements.

In order to be more broadly available to researchers, *seqr* has been made available as a connected application in the NHGRI’s Analysis Visualization and Informatics Lab-space (AnVIL). AnVIL is an application powered by Terra - a scalable and secure cloud-based platform for biological research. As a connected application, *seqr* legally resides within Terra’s security boundary. This means that users can be assured that the security controls around their data in Terra extend to *seqr* as well. Terra and AnVIL users can bring their own joint called VCFs, or create one using supported Terra workflows, and then load that data to their own private *seqr* projects for analysis. Due to its intended use as a research system, *seqr* does not currently have data fields for entry of Personally identifiable information (PII) or Protected Health Information (PHI). There are no restrictions for who can create an AnVIL account and then store data in their own private GCP bucket, which empowers researchers from a variety of backgrounds and technical abilities to access the *seqr* platform.

In addition to being available to all AnVIL users, the *seqr* platform is available as an open source project. Institutions or laboratories that prefer to operate their own *seqr* installation and have the technical resources to support it are invited to do so. GitHub is used to host the *seqr* code base as a public repository (https://github.com/broadinstitute/seqr) and is the same repository used by the Broad’s *seqr* deployment. Copyleft protection for *seqr* is licensed under the GNU Affero General Public License v3.0. Instructions for deployment are available in GitHub (see Supplementary Methods).

In addition to hosting *seqr’s* codebase, GitHub is also used for managing *seqr* technical requests. Bug reports and feature requests can be submitted via the repository’s “Issues’’. For questions and general *seqr* discussion, users are invited to join the *seqr* user forum (https://github.com/broadinstitute/seqr/discussions). In addition, *seqr* accepts code contribution for features and bug fixes from external groups via pull requests to the repository. We recommend users to first submit an issue to discuss such contributions with the *seqr* development team before starting on implementation.

## 8. Collaboration

Collaboration is a key feature of *seqr*. For example, our Broad CMG analysts work closely with over 50 collaborating research teams worldwide to diagnose cases by enabling groups to access a common workspace, tag candidate variants for group discussion, and jointly contribute to analyses. In addition, Broad CMG analysts search for candidate genes across multiple projects for potential matches and connect the respective research groups. This collaborative approach has empowered numerous diagnoses and novel gene discoveries across the more than 16,000 families we have studied to date (Coppens et al., 2021; Donkervoort et al., 2019; Mohassel et al., 2021). Due to joint-calling, the allele count across the Broad CMG callset is displayed for all variants in the search results and aggregation of cases with common etiologies is easily enabled. For example, analysts at Broad identified three other cases with the same homozygous splice variant in *TRAPPC4*, which led to its association as a relatively common cause of early-infantile neurodegenerative syndrome (Supplementary Figure 5) (Ghosh et al., 2021).

Both the Broad and University of Washington centers participating in the recently launched NHGRI Genomic Research to Elucidate the Genetics of Rare Disease (GREGoR) consortium are using *seqr* as their primary analysis and project management platform. Some of the sites using *seqr* outside of GREGoR include the Bahrain Genome Project, Boston Children’s Hospital (Boston, MA, USA), Garvan Institute of Medical Research (Sydney, Australia), Murdoch Children’s Research Institute (MCRI; Melbourne, Australia), National Institute for Allergy and Infectious Disease (Bethesda, MD, USA), Yale University (New Haven, CT, USA), University of Tartu (Tartu, Estonia), and University of Southampton (Southampton, UK).

New feature ideas and feedback on existing features often come from groups that use *seqr* through CMG collaborations. They submit feedback through *seqr’s* GitHub page which has led to many improvements in *seqr*’s analysis capabilities. In other cases, outside developers have added features needed by a local group using *seqr* and we have integrated these into the primary version of *seqr* so these features can be accessed by all users. An example of this is the recent integration of code for Genomics England’s and Australia’s PanelApp tool, making >300 curated disease panels now selectable in *seqr*. Other new features in development from external groups include the addition of filtration by in silico scores and the integration of a module to allow application of the ACMG/AMP criteria codes (Richards et al., 2015) for classifying variants according to pathogenicity. From regular discussions between analysts and developers, the variant search interface undergoes continuous iterative enhancements to improve the variant search experience.

## 9. Discussion

The *seqr* platform was designed to be a resource for the global rare disease research community, enabling efficient, high-quality analysis and collaboration. The main feature driving its utility is the user-friendly interface which allows quick onboarding to the platform by researchers and clinicians who are familiar with disease architecture and patient phenotypes but lack a computational background. The recommendation to use pre-populated standardized searches allows for consistency and reproducibility in the variant filtering and prioritization process across teams, while also allowing adjustment as needed. Recent development efforts have focused on incorporating additional data types (e.g. SVs from the GATK-SV pipeline) and providing support and guidance materials for data loading from different file formats from external users. With the addition of CNV calls from exome data with the gCNV caller, we have been able to make dozens of diagnoses of *de novo* CNVs, homozygous CNVs with recessive inheritance, and compound heterozygous variants, most often with a CNV in *trans* with an SNV or indel. It is in these cases where analysis is needed across the CNV and short variant data that *seqr* is particularly powerful for analysis. A popular feature of *seqr* is the ability to directly view the raw read data supporting a specific variant through a web-IGV integration. Future planned visualization integrations include support of RNA sequencing data to interrogate changes in expression and splicing.

Project management features have given users the ability to efficiently track the status of samples and cohort analyzed in *seqr*, and facilitated sharing of metadata in standardized formats for AnVIL as well as submitting candidate genes through the Matchmaker Exchange. And by enabling several variant types in *seqr*, users can have all project results in a single location. In cases where a causal variant is identified through an independent method, but is missing or inaccurate in the short-read variant calls stored in *seqr*, such as many short tandem repeat (STR) expansions, there is also an option for users to manually add the variant to *seqr* to ensure these diagnoses are recorded.

Seamless real-time integration with the Matchmaker Exchange platform, and detailed support for communication, has been a critical addition to *seqr*, given that most novel gene discoveries made by the Broad CMG have benefited from matches identified through MME. We continue to expand our interactions as new nodes are added to the federated MME platform.

Initially, *seqr* was designed to meet the project management, collaborative analysis, and data sharing needs of the Broad CMG. Local instances can be deployed but require computational expertise for installation and maintenance. To enable more widespread access to users without software engineering skills, *seqr* is now available on the AnVIL platform. Anyone can upload a joint-called VCF file into a private workspace in Terra or generate a joint called VCF from data available in AnVIL and then request the joint VCF be loaded in *seqr*. To further empower the global rare disease community, we have made written and video-based materials and training workshops materials available (https://cmg.broadinstitute.org/using-seqr).

New projects and new users will want additional features to be implemented in seqr, and we look forward to continuing to work with developers to improve and expand the platform. The recent Panel App gene list integration from the team at MCRI is a successful example of how features built to support a local use case can benefit all *seqr* users.

In summary, many platforms currently exist to support rare disease case analysis; however, most are expensive commercial platforms or proprietary and confined in use to a single laboratory. Access to a free and publicly accessible platform that enables collaborative analysis and communal code contribution is critical to enabling widespread equitable involvement in rare disease diagnosis and gene discovery as well as harnessing the latest methodological advances in genome analysis. Engaging the larger rare disease community to contribute to developing *seqr* and increasing the rare disease genomic data that can be shared within *seqr* or easily exported to other platforms will advance rare disease science as well as patient diagnosis and treatment.

## Data Availability

The *seqr* platform code is available at https://github.com/broadinstitute/seqr. Training videos are available at https://cmg.broadinstitute.org/using-seqr. Candidate genes identified by the Broad Institute Center for Mendelian Genomics (Broad CMG) have been submitted through *seqr* to the Matchmaker Exchange
(https://www.matchmakerexchange.org/) and shared through the CMG website (mendelian.org). De-identified and coded genomic and phenotype data have been shared on the National Human Genome Research Institute (NHGRI) AnVIL platform (https://anvilproject.org/data). Data access requests can be made per instructions here https://anvilproject.org/learn/accessing-data/requesting-data-access#accessing-controlled-access-data or can be obtained through dbGaP (study ID phs001272).

http://mendelian.org/phenotypes-genes

https://anvilproject.org/data

https://github.com/broadinstitute/seqr

https://cmg.broadinstitute.org/using-seqr

## Acknowledgements

We thank Dr. Simon Sadedin, David Ma and Tommy Li for their contributions to integrating PanelApp into *seqr*; and Brett Thomas for his contributions to the development of an early precursor to the *seqr* platform. Platform development was supported by the National Human Genome Research Institute (NHGRI), the National Eye Institute, and the National Heart, Lung and Blood Institute grant UM1HG008900, NHGRI grants R01HG009141, U24HG010262, and U01HG011755.

## Ethical Considerations

This project has been reviewed and approved by the Mass General Brigham Institutional Review Board.

Variant data are based on real cases, but individual data in the manuscript (including sample IDs, names, email addresses, ages, phenotypes and ancestry) were created for illustrative purposes or edited to protect patient confidentiality.

## Data and Code Availability Statement

The *seqr* platform code is available at https://github.com/broadinstitute/seqr. Training videos are available at https://cmg.broadinstitute.org/using-seqr. Candidate genes identified by the Broad Institute Center for Mendelian Genomics (Broad CMG) have been submitted through *seqr* to the Matchmaker Exchange (https://www.matchmakerexchange.org/) and shared through the CMG website (mendelian.org). De-identified and coded genomic and phenotype data have been shared on the National Human Genome Research Institute (NHGRI) AnVIL platform (https://anvilproject.org/data). Data access requests can be made per instructions here https://anvilproject.org/learn/accessing-data/requesting-data-access#accessing-controlled-access-data or can be obtained through dbGaP (study ID phs001272).

## Web Resources

1. https://anvilproject.org/
2. http://care4rare.ca/
3. http://compbio.cs.toronto.edu/silva/
4. http://mendelian.org/
5. http://www.informatics.jax.org/
6. https://anvilproject.org/
7. https://clinicalgenome.org/
8. https://gemini.readthedocs.io/en/latest/
9. https://genome.ucsc.edu/
10. https://gnomad.broadinstitute.org/
11. https://gtexportal.org/home/
12. https://igv.org/
13. https://iobio.io/
14. https://monarchinitiative.org/
15. https://omim.org/
16. https://panelapp.genomicsengland.co.uk/
17. https://phenodb.org/
18. https://www.deciphergenomics.org/
19. https://www.matchmakerexchange.org/
20. https://www.mousephenotype.org/
21. https://www.ncbi.nlm.nih.gov/clinvar/
22. https://www.sanger.ac.uk/tool/exomiser/

## Author Contributions

LSP contributed to the development of the platform and wrote the manuscript. HS developed the platform and contributed to the manuscript. DGM, HLR and AOL contributed to conceptualization and development of the platform and editing the manuscript. All other authors contributed to developing the *seqr* platform and reviewing the manuscript.

