## Supplemental Methods and Data for "*seqr* : a web-based analysis and collaboration tool for rare disease genomics"

### Supplementary Data

#### Frameworks

The *seqr* web application is built using Django, a Python web framework, to handle server-side functionality such as URL routing and database querying via its Object Relational Mapper (ORM). For the front end, *seqr* leverages React to provide an extensible, component-based user interface, integrated with Redux to handle state management. Semantic UI (via the Semantic UI React package) is used for Cascading Style Sheets (CSS) to ensure consistent styling throughout the application. While the *seqr* application is designed to be platform agnostic, it is currently hosted on Google Cloud Platform (GCP). Kubernetes is used to manage deployment and resource management within GCP, including using kubernetes secrets to handle sensitive deployment parameters.

#### Databases

Data storage in *seqr* is separated into two different databases: a read-only database which includes all annotated variant information, and a read/write database which includes all project and user metadata, including user-generated data like variant tags and notes.

For the variant database, *seqr* leverages Elasticsearch (ES), a powerful no-SQL database optimized for flexible data queries. Data is loaded via an external pipeline, and *seqr* accesses the data in a read-only capacity. This ensures data integrity against malicious or accidental user manipulation. The data loading pipeline (https://github.com/broadinstitute/hail-elasticsearch-pipelines) utilizes Hail to parse, annotate, and format a joint-called VCF with the metadata required for search and display in *seqr*. These annotations include population frequencies, in silico predictor scores, VEP consequences, and sample-specific VCF data such as quality metrics and zygosity. This data is then exported into project-specific ES indices, and indices are never modified after their creation. When new or updated data becomes available, data is re-exported to a new index and the old index is deleted.

All other application data is stored in postgres databases. The primary postgres database contains all user-provided application data. This includes user account information, metadata about projects, cases, and samples, user-generated gene lists and search criteria, and saved variant information. User-provided variant information includes tags, notes, and variant annotations such as transcript of interest. In addition, a copy of any variant saved by users in *seqr* is stored in postgres as well as Elasticsearch in order to improve application performance and consistency. Additionally, *seqr* stores external reference data in a separate postgres database. Such reference data includes gene level information from sources such as GENCODE, OMIM, and dbNSFP, as well as structured phenotype data from HPO. This reference data is updated periodically to ensure accuracy but is not editable by *seqr* application users. Additionally, *seqr* leverages redis for caching key request data in order to optimize application performance.

### Security

As a connected application, *seqr* legally resides within Terra's security boundary and all security controls around the data in Terra extend to *seqr* as well. In order to mitigate potential data breaches, *seqr* operates as a FISMA Moderate system (Federal Information Security Modernization Act; https://www.cisa.gov/federal-information-security-modernization-act) and is compliant with the NIST 800-53 Rev 4 information security standard at the Moderate baseline.

### Local installations

In addition to being available to all AnVIL users, the *seqr* platform is available as an open source project. Institutions or laboratories that prefer to operate their own *seqr* installation and have the technical resources to support it are invited to do so. GitHub is used to host the *seqr* code base as a public repository (https://github.com/broadinstitute/seqr) and is the same repository used by the Broad's *seqr* deployment. Copyleft protection for *seqr* is licensed under the GNU Affero General Public License v3.0 (https://github.com/broadinstitute/seqr/blob/master/LICENSE.txt). Docker and docker compose are used to manage pre-built *seqr* resources to streamline deployment. Instructions for installation onto any cloud platform or on-prem server are available in the GitHub repository (https://github.com/broadinstitute/seqr/blob/master/deploy/LOCAL_INSTALL.md).

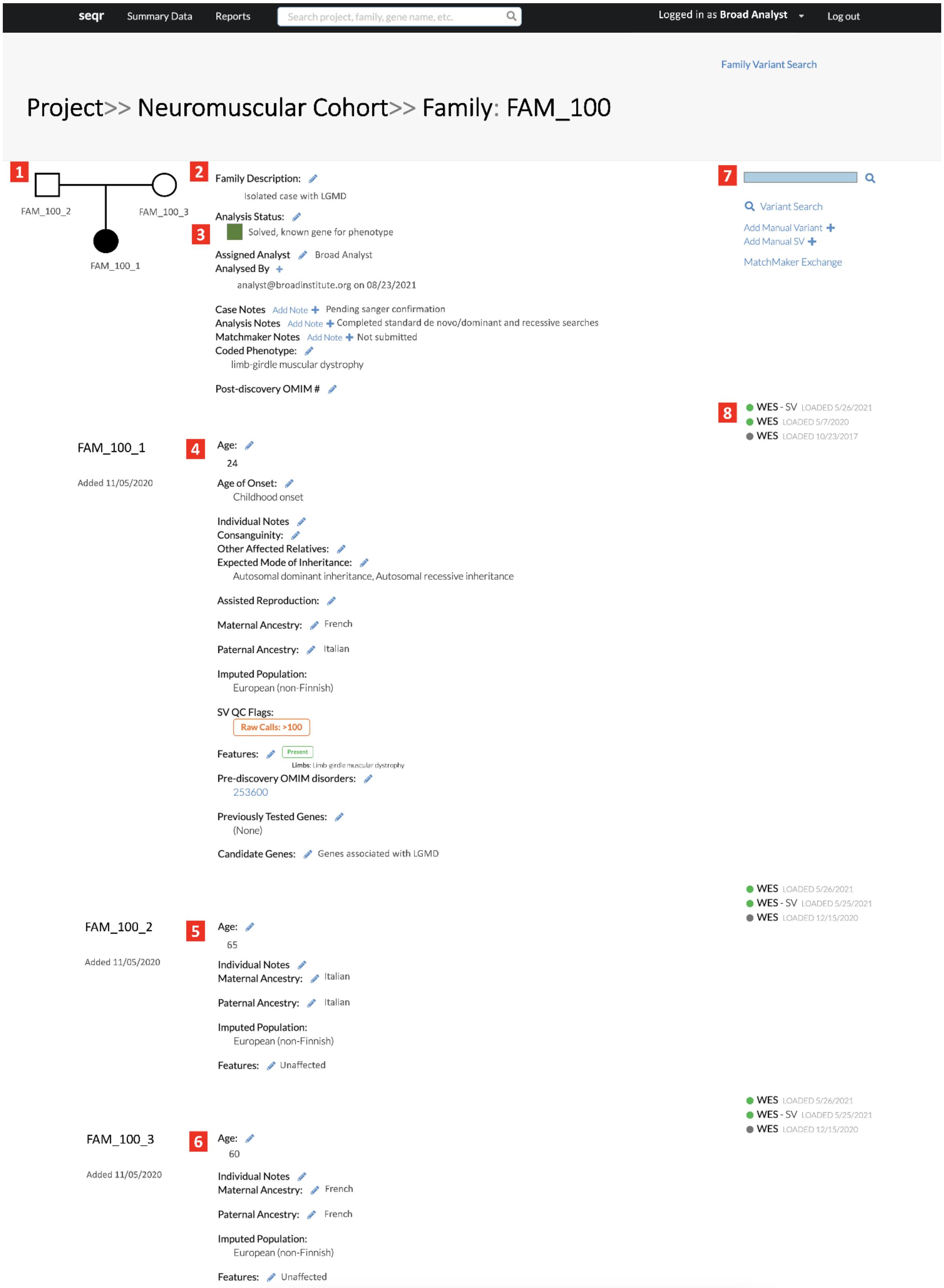


**Supplementary Figure S1:** The *seqr* Family Page displaying FAM_100 as an example**.** (1) family pedigree; (2) general description of the case; (3) analysis and case details entered by the analysis team; (4-6) individual level example data including age, ancestry, sample QC information, clinical information using HPO terms; (7) variant search link to begin analysis and an overview of tagged variants on the hover over of the saved variants box, if the case was previously analyzed; (8) type and date of data loaded into *seqr*.

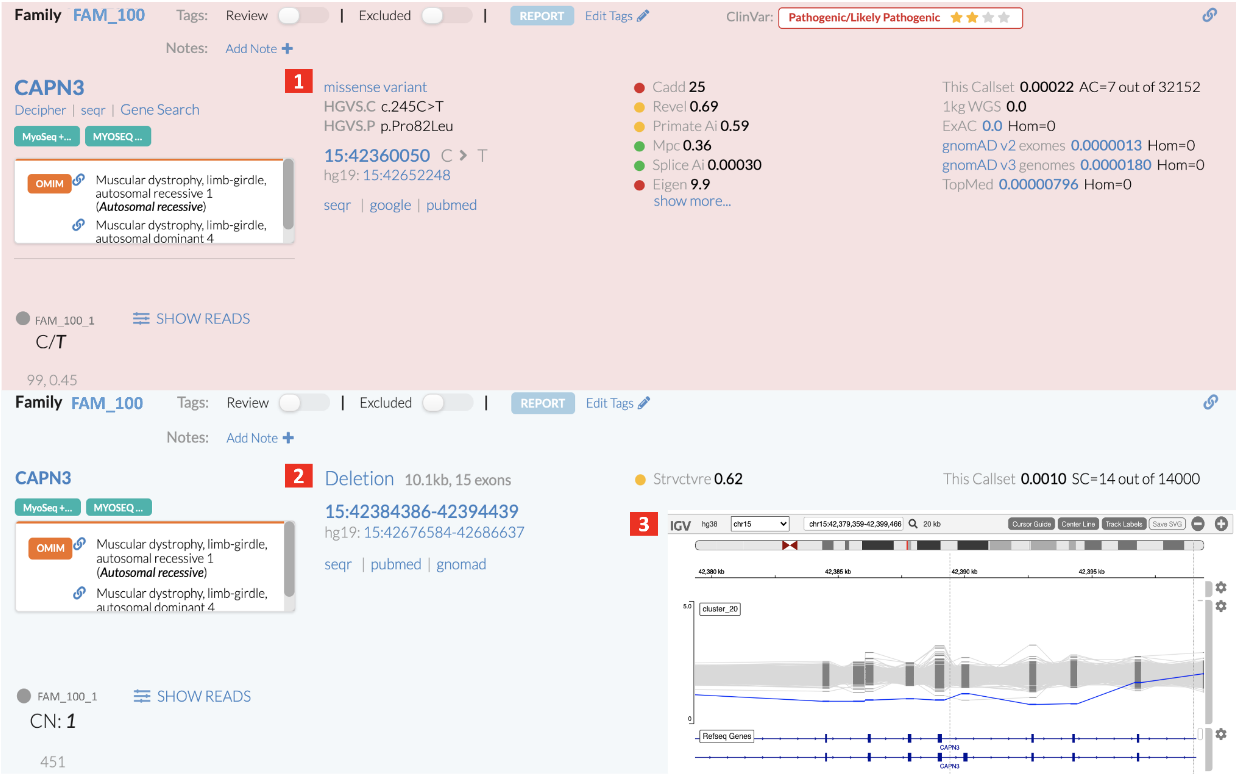


**Supplementary Figure S2:** Variant filtration of SNVs/indels and SVs in tandem. In this example, a recessive restrictive search identified (1) a missense variant and (2) deletion in *CAPN3.* Variants in the raw read data can be viewed using (3) IGV within *seqr*. Both variants were externally validated and reported as the diagnosis for this research participant with limb-girdle muscular dystrophy.

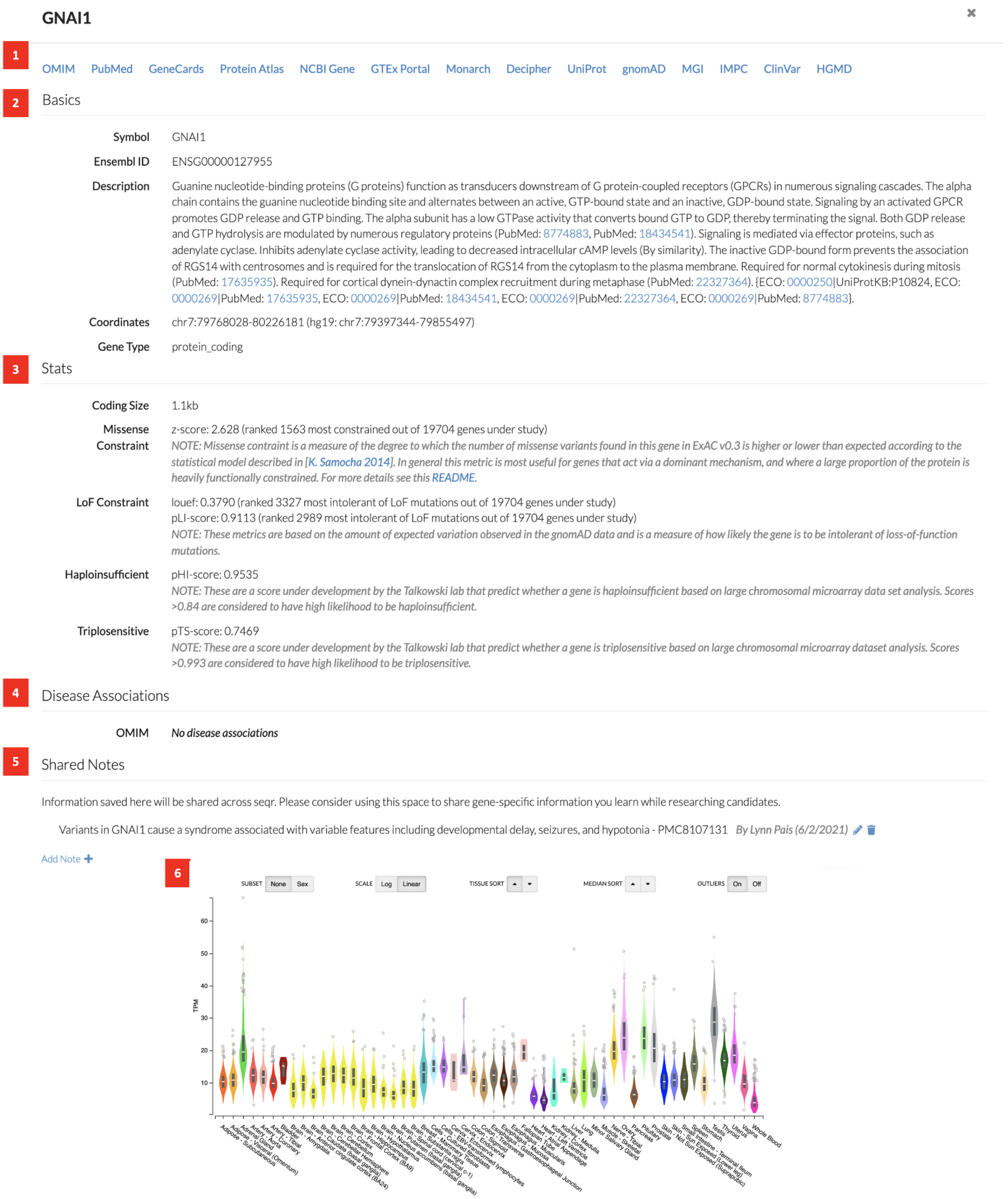


**Supplementary Figure S3:** Gene Page. Includes (1) links to external resources; (2) basic gene information; (3) gene constraint and dosage sensitivity details; (4) gene-disease relationships based on OMIM; (5) user notes shared across *seqr*; (6) GTEx tissue expression data.

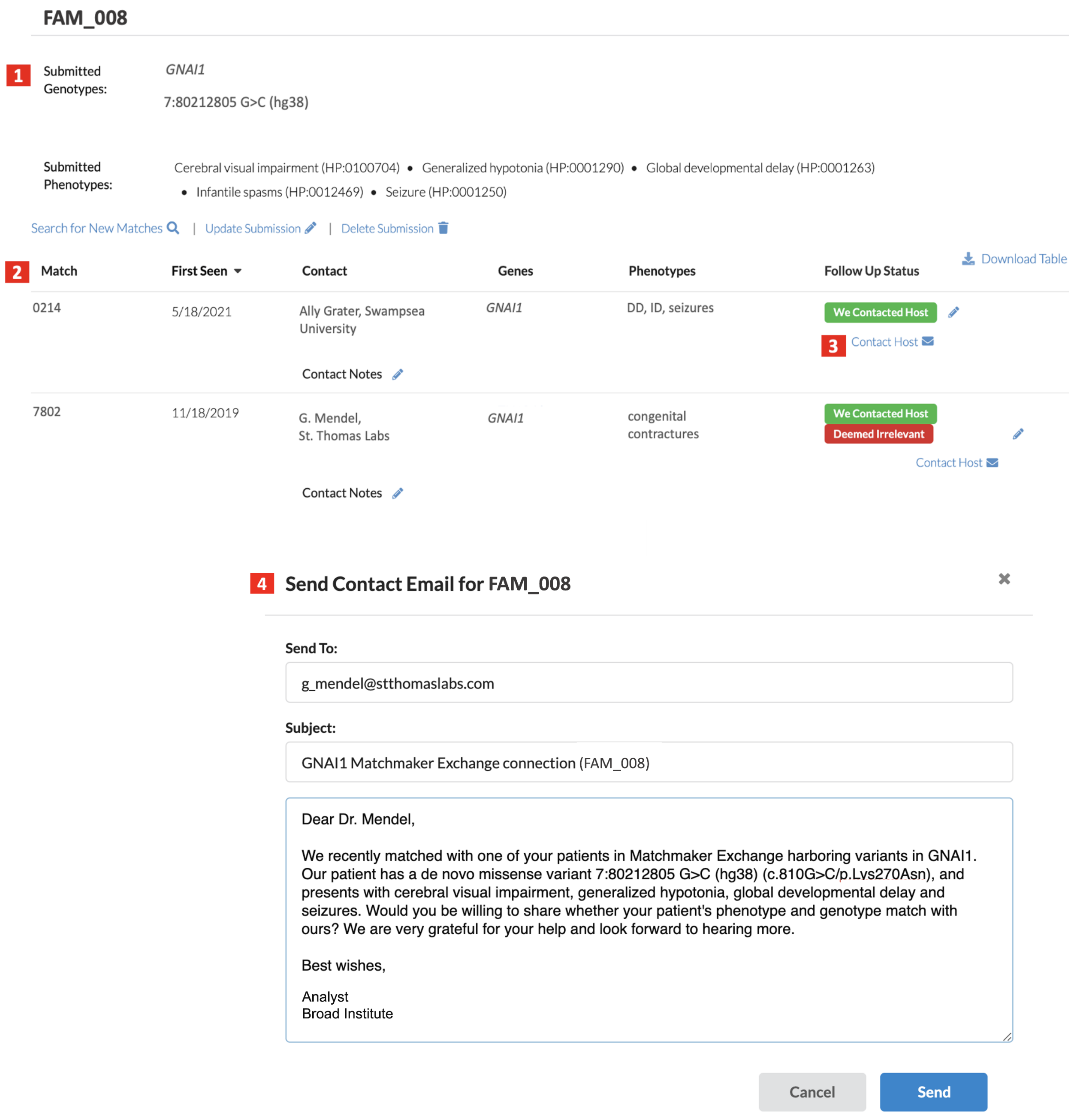


**Supplementary Figure S4:** The Matchmaker Exchange (MME) node in *seqr*. (1) variant and HPO terms listed in *seqr* that were submitted to MME; (2) examples of MME matches with host's contact details, gene ID, and phenotype, if included in the submission; (3) contact feature to communicate with hosts and track the status of matches within *seqr*; (4) autogenerated email with sample variant and phenotype information from *seqr*.

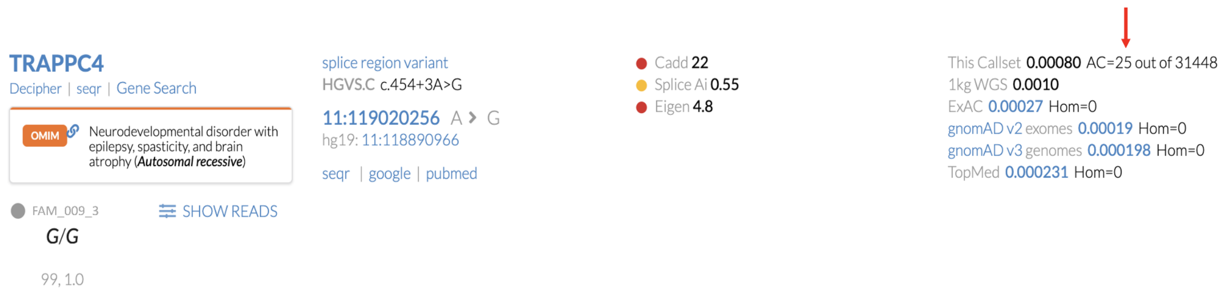


**Supplementary Figure S5:** Allele count for a splice region variant in the gene *TRAPPC4* showing 25 alleles in the CMG callset. A review of the variants revealed three other cases with similar phenotypes that were this homozygous for this variant across distinct projects and research groups.
